# How can we compare CI systems across manufacturers? A scoping review of recent literature

**DOI:** 10.1101/2023.06.13.23291338

**Authors:** Elinor Tzvi-Minker, Andreas Keck

**Affiliations:** Syte Institute, Hamburg, Germany

**Author notes:** Correspondence should be addressed to: Dr. Elinor Tzvi-Minker, Syte Institute, Hohe Bleichen 8, 20354, Hamburg, Germany.

**Keywords:** Cochlear implant, Patient-reported outcomes, Pure Tone Average, Speech in noise, Music perception

## Abstract

Electric stimulation via a Cochlear Implant (CI) enables people with severe to profound sensorineural hearing loss to regain speech understanding and music appreciation and thus allowing them to actively engage in social life. Three main manufacturers (Cochlear, MED-EL and Advanced Bionics “AB”) have been offering CI systems, thus challenging CI recipients and Otolaryngologists with a difficult decision, as currently no comprehensive overview or meta-analyses on performance outcome following CI implantation is available. The main goal of this scoping review is to provide evidence that data and standardized speech and music performance tests are available for performing such comparisons. To this end, a literature search was conducted to find studies that address speech and music outcomes in CI recipients. From a total of 1592 papers, 188 paper abstracts were analyzed and 147 articles were found suitable for examination of full text. From which, 42 studies were included for synthesis. A total of 16 studies used the consonant-nucleus-consonant (CNC) word recognition test in quiet at 60db SPL. We found that aside from technical comparisons, only very few publications compare speech outcomes across manufacturers of CI systems. Evidence suggests though, that these data are available in large CI centers in Germany and US. Future studies should therefore leverage large data cohorts to perform such comparisons that could provide critical evaluation criteria and assist both CI recipients and Otolaryngologists to make informed performance-based decisions.

## 1. Introduction

Cochlear implants (CI) encompass implanted electronics and external sound processors that can deliver electric stimulation to the auditory nerve and improve hearing in subjects with severe to profound hearing loss. Despite being a well-established intervention for this condition, there is a strong variability in the individual hearing restoration achieved by CI, which may depend on several factors, ranging from device specifications to surgical placement of the implant, as well as patient-specific factors such as age at implantation and duration of hearing loss (Holden et al., 2013). Implant recipients improve their conversational speech understanding following implantation, on average up to 52% (Dunn et al., 2020) and in many cases, improve so significantly that they can understand conversational speech in difficult listening situations (Dorman & Gifford, 2017). Significant improvement in music perception and satisfaction following CI implantation is also observed (Chang, 2021). This has become particularly important as studies showed that quality of musical sound and patients QoL post-implantation were associated significantly. However, CI were initially designed to enhance speech discrimination. In the past 10 years, Fine Structure information has been represented in CI processing strategies to further improve music perception (Lassaletta et al., 2009).

Recent estimations suggest that approx. one million cochlear implants have been implanted world-wide (Zeng, 2022). Clearly, a rapid growth in this field can be observed as previous FDA (food and drug administration, USA) reports from 2019 and 2016 suggest an approx. of 736K and 324K resp. implanted devices worldwide. A portion of this growth can be attributed to expanding CI candidacy guidelines (Varadarajan et al., 2021). Implantation is now available to a broader group of individuals compared to when implants were first introduced in the 70’s. Individuals are now more commonly implanted with a CI system when they suffer from unilateral deafness (Carlson et al., 2018), or intractable tinnitus (Assouly et al., 2021), and in the presence of increasing amounts of residual hearing (Varadarajan et al., 2020).

Accordingly, the number of scientific studies in the field of CI has increased exponentially as implants have become widely available and candidacy guidelines have expanded. A recent search for cochlear implants on PubMed yielded over 1500 papers in 2021 alone, an increase of almost 600% in the last 20 years (2001: 262 papers). Study topics in peer-review literature range widely, from candidacy guidelines to implant technological features, programming (mapping), rehabilitation, and performance outcomes to name a few.

However, less attention has been devoted to comparative performance of CI users of different commercially available technologies, although four main manufacturers, Cochlear, Advanced Bionics (AB), Oticon Medical (previously Neurelec and recently acquired by Cochlear) and MED-EL, have been offering CI systems since the 1990’s. A group of CI users who are implanted with CI of different brands in each ear, are able to compare sound quality and performance, however these subjects are rare. A study by Harris et al, (2011) evaluated speech and music perception through two different brands of CI in the same subject. Six subjects were implanted with a Cochlear Nucleus in one ear and subsequently implanted with a MED-EL Sonata in the contralateral ear. While no difference in perception was seen in objective testing, subjective music perception was found to be superior with the MED-EL implant in most subjects. This evidence coincides with anecdotal testimonials from CI-carriers who performed revision surgery to a CI by MEDEL^1^. Thus, performance comparisons between brands are important and have a strong effect on quality of life of CI users.

While clearly the differences between CI systems includes specific technological features, in this scoping review, we aim to highlight the difference in performance outcomes following CI implantation of systems by different manufacturers. This is an aspect that is usually overlooked in comparative studies and should gain more focus. Indeed, in the last decade, patient-reported outcomes (PRO) have gained importance in guiding advancements in medical technology and influence healthcare policy and practice (Deshpande et al., 2011). A recent review by Stucke et al. (MedRxiv)^2^ focusing on PRO of implanted devices found 47 studies in the last 22 years that report on PRO for CI, including speech and music perception outcomes. However, only 17 studies out of these 47 provided any information on device manufacturers. This means that the data needed to compare speech and music performance outcomes between the different devices apparently exist, but comparisons are seldomly made.

One outstanding example, dating 15 years back, is a study by Spahr et al. (2007) which investigated patient’s performance in monosyllabic word tests presented in quiet and under different noise levels, comparing CI systems by the three manufacturers (Cochlear, AB and MED-EL). Results showed differences between devices in vowel recognition, and sentence comprehension in noise. In addition, the authors showed that when the input dynamic range was increased, performance measures of all devices improved. A later study by Haumann et al. (2010) compared speech performance in noise under realistic adaptive conditions across five different CI systems (Freedom and Esprit 3G by Cochlear, Auria and Harmony by AB, Opus 2 by MED-EL). Here results showed a clear disadvantage for Freedom (Cochlear) compared to Opus 2 (MED-EL). Other studies have compared specific technical features of these systems (Shader et al., 2020; Sivonen et al., 2020). For example, Killan et al. (2019) assessed the effect of inter implant interval and onset of profound deafness on sound-source localization in children with bilateral cochlear implants from three manufacturers (Cochlear, AB and MED-EL). The authors found that MED-EL devices were associated with significantly better sound-source localization when compared to both Cochlear and AB devices.

A recent retrospective study by Sturm et al., (2021), investigated the effect of physical features of CI electrodes including length and shape of the electrode array, as well as its position relative to the cochlear modiolus, on hearing outcomes. Authors recruited 119 adult (>18 years) subjects with post-lingual hearing loss, who underwent cochlear implantation with a full electrode array insertion. Seven different electrode arrays from three CI manufacturers (Cochlear, MED-EL, AB) were compared. Speech perception outcomes were measured using the consonant-nucleus-consonant (CNC) word recognition test in quiet, at the same presentation level and fixed test intervals (three, six, 12, and 24-months following implantation). Pre-operative speech scores were similar between electrode array groups and the same surgical approach was used. Given the consistencies in data collection and patient demographics, this study was well configured for a comparison between the devices. However, the authors chose to compare speech performance without accounting for pre-operative measures of CNC and thus dramatically increased inter-individual variability as well as reduced reliability of device comparisons. In addition to the pre-operative speech perception score, several other factors should have been accounted for such as: (1) The duration of deafness and pre-operative hearing aid used, and (2) the cochlear duct length, insertion angle and electrode position within the cochlea. Lastly, while CNC in quiet may provide some insights on hearing abilities, other speech tests performed in noise and thus assimilating real-life scenarios, are much more suitable for assessing the range of auditory abilities following implantation.

A review by Boisvert et al. (2020) further supports the possibility to conduct comparative studies across CI manufacturers. The authors aimed to provide evidence for the efficacy of unilateral cochlear implantation in adults by assessing the procedure’s success rate, based on speech perception or self-reported measures in studies from the last 22 years. The authors found that while measurements, research design and reporting of results were inconsistent, 46 studies used monosyllabic words for post-operative speech perception tests, while 34 studies used sentences in quiet to test for speech perception following CI implantation. In addition, there was some relative consistency with presentation levels of monosyllabic words in quiet, with mostly being 60dB SPL (32% of studies) or 65dB SPL (36% of studies). Such numbers suggest a potential for a meaningful comparison of speech outcomes between devices of different manufacturers.

To be able to compare data from existing publications and determine if speech and music performance outcome using a specific CI system is superior, several factors need to be considered. Firstly, it would be important to compare devices of the same generation. As CI technology advances, patient performance improves (Krueger et al., 2008). Therefore, a comparison of outcomes with the latest technology to previous generation devices is not sensible. Another consideration is the test conditions. Ideally, identical conditions are necessary to systematically compare performance outcomes. Evaluation tools, presentation levels, signal to noise ratios, language and test intervals would need to be similar if not identical to be able to perform meaningful comparisons (Adunka et al., 2018). Lastly, subject demographics must be considered. Duration of deafness, prior use of amplification, pre-operative pure tone thresholds (PTA), and certain etiologies are all known factors that may impact speech performance following implantation (Ahmed Elkayal et al., 2016). Finally, similar patient profiles would be important in isolating the effect of the CI system in driving outcome differences and therefore allow optimal comparison between CI systems of different manufacturers.

The main goal of this scoping review is to identify a research gap in comparisons of speech and music perception outcomes following CI implantation across different manufacturers. Manufacturer comparisons focused on outcomes are of crucial importance for clinicians, CI candidates and manufacturers. Such comparisons could be an important, more straightforward, and reliable source for decision-making processes, when compared to various technical device features that differ between CI systems^3^. CI manufacturers could benefit from this transparency by better understanding the effect of technological advancements on patient outcomes and factor these key learnings into future developments. Evidence suggests that more competitive markets within the healthcare industry, lead to increased quality of product features (Medicine et al., 2010).

## 2. Methods

A scoping review methodology was chosen to evaluate whether speech and music perception outcomes in adult CI users could be compared between manufacturers. A scoping review is ideal for answering these types of questions, by providing a coverage of a body of literature on a given topic, thus giving a clear indication of the availability of studies (Munn et al., 2018). We applied the population, concept, context (PCC) framework recommended for scoping reviews which guided the protocol listed below. The population is hearing impaired adults who underwent implantation of CI system. The concept of the scoping review is speech and music perception outcomes, and the context is defined to be the availability of CI manufacturer information.

### 2.1 Search strategies

A literature search was conducted using both PubMed database and Google Scholar search engine, thus covering a broad literature source for the field of Cochlear Implants. We used the key words “Cochlear implant outcomes adults” (S1) as well as “Cochlear implant music adults” (S2) in March 2022. The review protocol was not pre-registered. An initial search found that Google Scholar showed less relevant publication titles when compared to PubMed and the publications found in the former matched those found on PubMed. In addition, we searched the clinicaltrials.gov database, which provides information on funded clinical trials around the world, with the search term: “Cochlear Implant” (S3) in March 2022. We cross-checked the findings with the first 200 papers on Google Scholar. Fig. 1 describes our screening procedure.

**Figure 1.**
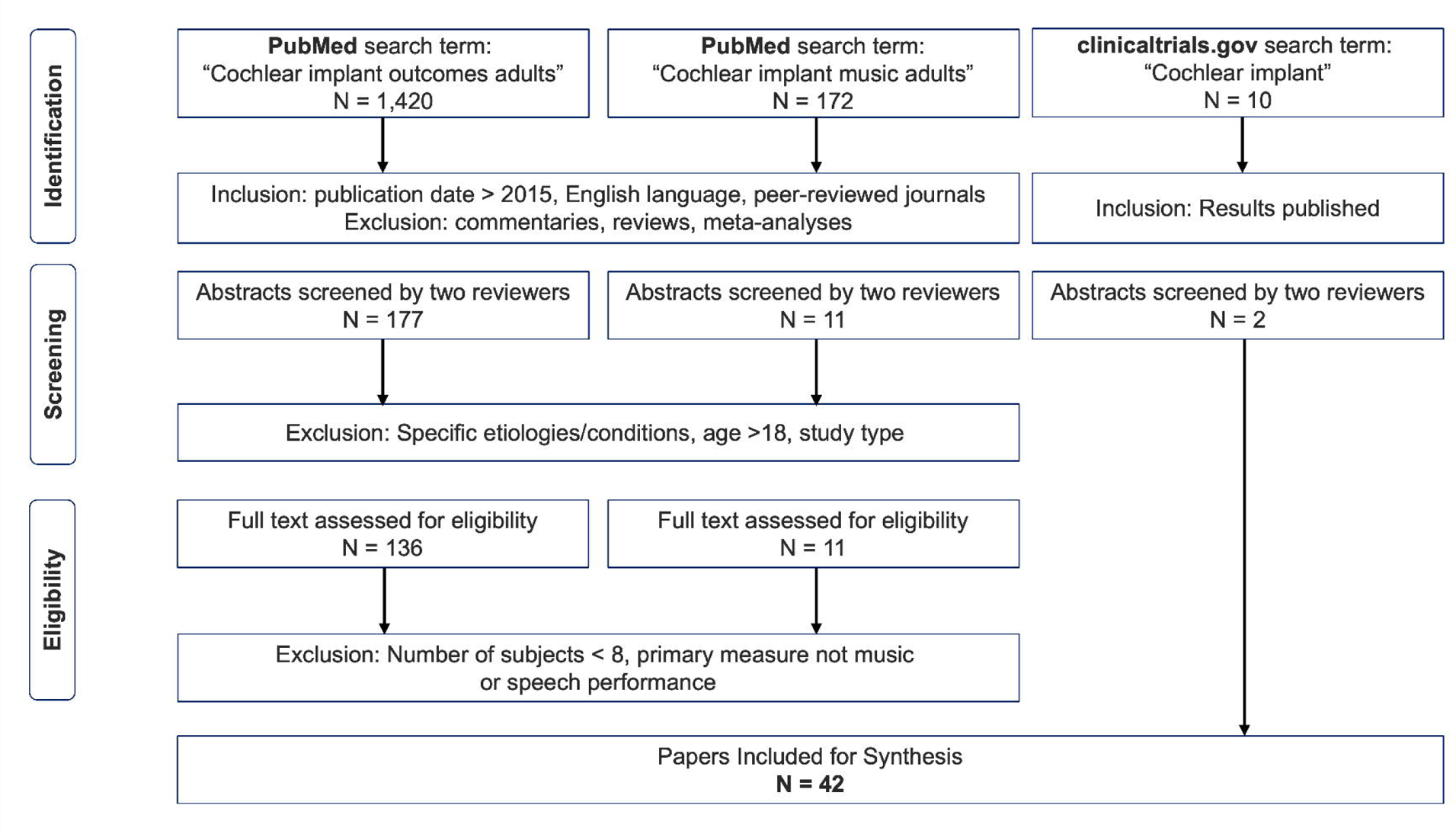
Literature Inclusion Flowchart

### 2.2 Eligibility criteria

The authors independently selected all English-language, peer-reviewed studies published after 2015. This time frame was selected to include speech and/or music perception outcomes of latest technology, as comparisons to previous generation devices is not sensible (see introduction for further explanation). This resulted in 1,420 papers for S1 and 172 papers for S2. The authors then discarded all reviews, commentary articles, case-studies and meta-analyses as these do not contain detailed information required for our scoping review (mainly speech and music perception scores and manufacturer information). Next, the abstracts of all remaining studies (S1 = 177, S2 = 11) were analyzed independently and charted in Excel (© MS Office). The authors applied the following exclusion criteria to the remaining studies through a mutual discussion.

- Exclusion based on indication Reports on populations with the same etiologies/conditions, e.g., Meniere’s disease, auditory deprivation, vestibular schwanomas, active military duty, prelingually deaf or cognitive decline were excluded as these factors are known to impact performance
- Exclusion based on participant-specific criteria Subjects <18 years of age
- Exclusion based on study type Studies focusing on predicting factors that influence performance outcome such as: genetics, fatigue, subject self-reports, candidacy, robotics, telemedicine, auditory training, surgical approaches, reimplantation, surgical complications, or revision surgery. These studies were excluded to eliminate the inclusion of very specific populations that may differentially affect performance which and are not representative of the CI community. Longitudinal studies were excluded as they include previous generation devices. Comparison of CI performance with other technologies (hearing aids, bone conduction implants). Drug therapies provided in addition to implantation. Objective measures not accompanied by speech scores. Listening effort or hearing preservation as a primary outcome measure.

Following this exclusion, a total of N = 147 (S1 = 136, S2 = 11) publications were found suitable for further analysis and an examination of the full text. We then additionally excluded studies with number of subjects lower than eight and primary measures that were not speech or music performance. Following this in-depth review of studies, we found 42 publications that we included in the final overview table (see supplementary materials).

### 2.3 Data charting process

A data-charting form was created in Excel by two reviewers (AK, ETM). Variables to be extracted from the studies were determined by all authors. Data was extracted as reported in the text or the figures. Study authors were not contacted when study information was unclear or not reported. Data charting categories included general information such as the publication title, authors name, institution in which data was collected, year and journal of publication. In addition, device-specific information was collected such as device type and launch date as well as manufacturer information. Note that no studies with bi-branded CI recipients were included. Study characteristics such as the number of participants included in the study, age range, gender, as well as pre-operative parameters such as pure tone average (PTA), and duration of hearing loss were recorded. Importantly, we charted detailed information on the specific post-operative speech or music performance test, including test conditions, such as whether it was performed in noise or not as well as test interval following CI implantation.

### 2.4 Synthesis of results

We mapped the findings based on the following criteria: (1) study characteristics, including publication year and journal, (2) manufacturer information and launch data (3) speech tests, (3) speech test conditions, (4) music performance, (5) number of subjects (histogram). Analysis of speech perception outcomes focused on the most used test: CNC word recognition. The data from six studies that used CNC words presented at 60dB SPL in quiet were further compared. Three of these studies reported on devices manufactured by Cochlear, and three on devices manufactured by MED-EL. To account for the different number of participants in each study included in this analysis, we performed a weighted averaging of the scores by the number of participants. We synthesized the results for different intervals including 1-month, 3-month, 6-month and 12-month following implant operation.

## 3. Results

### 3.1 Study Characteristics

The 42 studies included are detailed in the supplementary materials. In Fig. 2, we show the distribution of studies across 2015 - 2021. Most studies were published in 2020 (N = 9) and a relatively low number of studies were published in 2017 (N = 3) and 2021 (N = 4).

**Figure 2.**
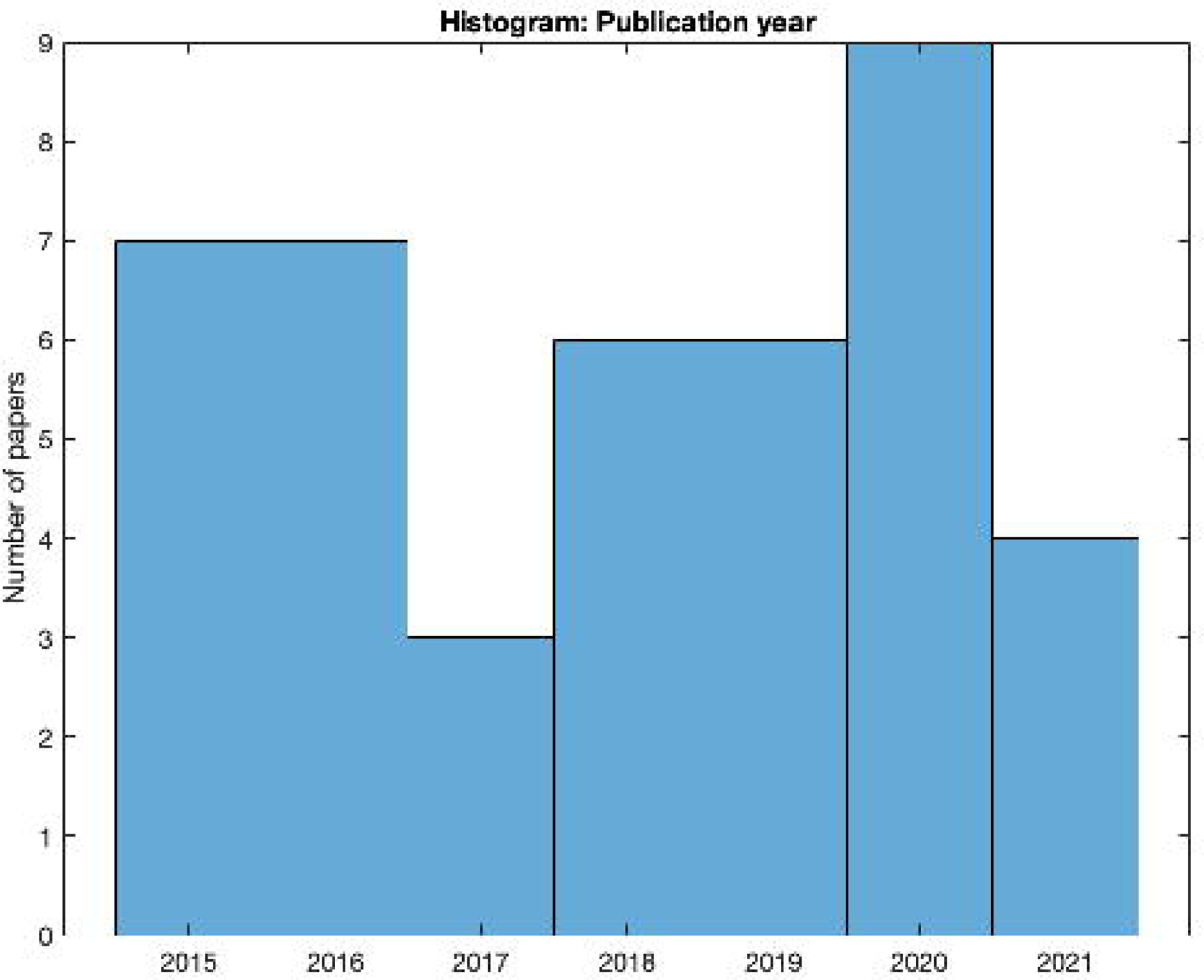
Number of studies published per year from 2015 to 2021

The distribution of journals used to publish speech and music perception outcomes for CI is depicted in Fig 3. Most studies were published in Otology & Neurotology (N = 9) followed by Cochlear Implants International (N = 6). Only three studies were published in JAMA Otolaryngol Head Neck Surg.

**Figure 3.**
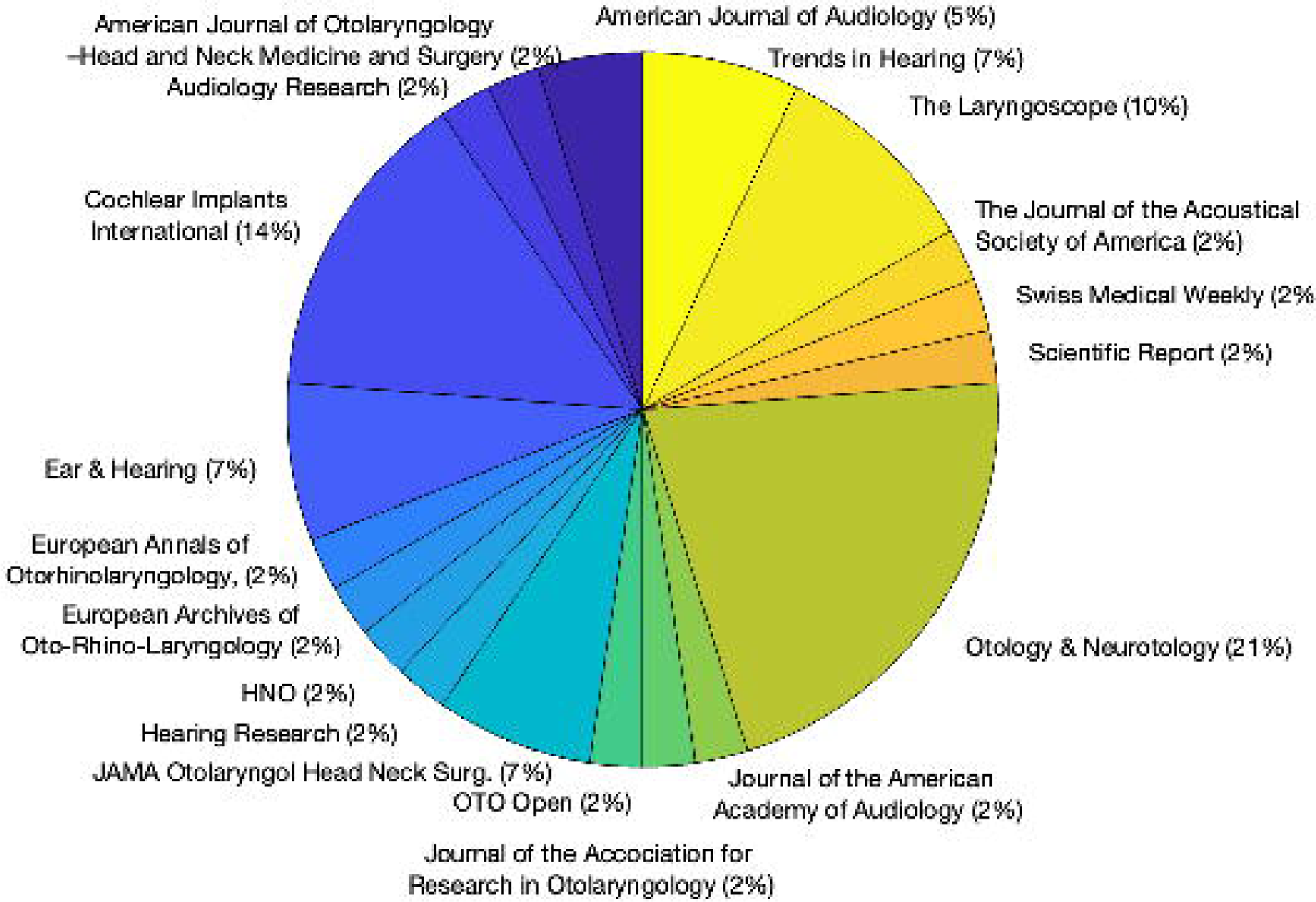
Distribution of Journals for the analyzed studies

### 3.2 CI system manufacturer information

Only two studies out of 42 did not mention any information regarding the manufacturer of the CI systems used in the study. From the remaining 40, five studies did not mention any information on the specific model of the CI system. The distribution of studies mentioning specific CI systems is shown in Fig. 4. Most studies mention Cochlear as a manufacturer for the CI systems implanted in the included cohort (N = 24). MED-EL is mentioned by 19 studies, AB by 12, and Oticon by three. Five studies mentioned three manufacturers, and 5 studies mentioned two manufacturers.

**Figure 4.**
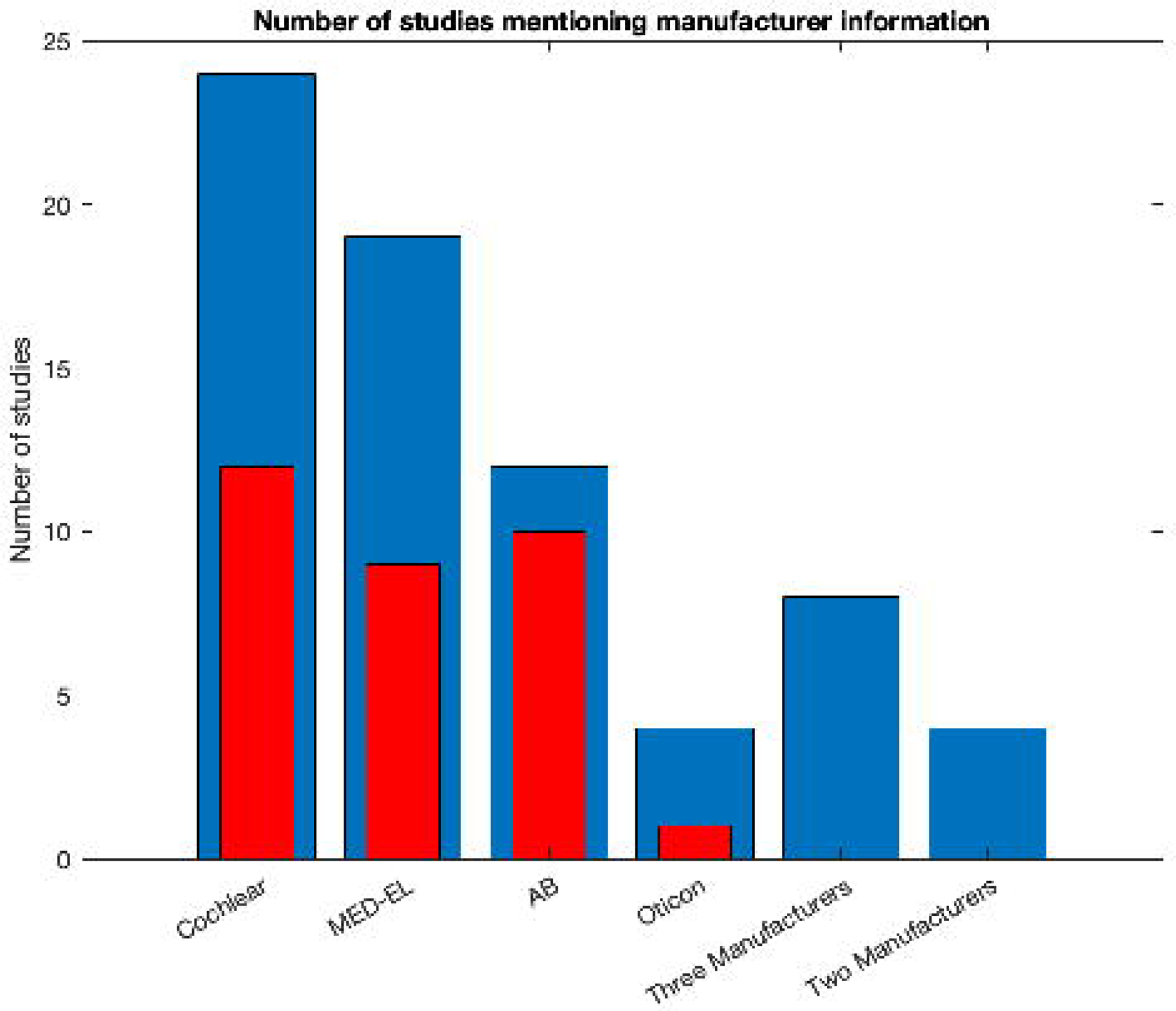
Blue bars show the number of studies identified for each CI manufacturer and when 2 or 3 manufactures were mentioned. The red bars show the proportion of studies of multiple manufactures mentioning a specific one.

### 3.3 CI systems launch date

Of the studies mentioning the manufacturer, we noted the device types to compare the date of paper publication with the novelty of the device technology. Only 24 studies (of 42, 57%) mentioned the device type. On average, CI systems were seven years old when the study was published. For studies including different manufacturers, the differences in launch dates were as much as 23 years (e.g., Fuller et al. 2021). Thus, direct comparisons in such studies were deemed invalid.

### 3.4 Sample size and a post-hoc power analysis

The total number of subjects ranged from eight to 2247, with two studies having less than 10 subjects investigated. Most studies had between 10 to 20 subjects investigated (See Fig. 5). Two of the 42 studies we investigated provided a power analysis to estimate the sample size needed. Neben et al., (2018) evaluated the number of subjects needed for speech perception performance test in noise based on a previous Hybrid Hearing study (Lenarz et al., 2013) that determined a 1.1dB difference to be clinically relevant. They found that 20 subjects are needed to reach a power of 83% for within-subject comparisons. The other study by (Williges et al., 2019) estimated an effect size of 0.56 for bimodal CI users that would require 21 subjects for within-group comparisons at 80% power. However, for conducting between-group comparisons to assess speech performance differences between CI systems of different manufacturers, a between-group comparison is required. Here we estimated how many patients would be needed to make valid comparisons between CI manufacturers, based on a statistical power analysis of results from (Sturm et al., 2021). These estimates could guide future studies that wish to perform comparisons within one or two CI centers but are concerned with the statistical power of such comparisons. Significant differences in CNC score were found at 3 and at 6 months post-implantation using standard t-tests. Given the data (means and standard deviations) provided in the paper, and assuming a power of 95% and alpha of p = 0.05, we found an effect size of 1.08 for the 3-month comparison and 1.04 for the 6-month comparison. Based on these rather large effect sizes, we estimated that a minimum total sample size of 42 subjects will be needed to find significant effects, when using a parametric Wilcoxon-Mann-Whitney test for differences between two groups (i.e., two manufacturers). The analysis was performed using G*Power (Faul et al., 2007). From the studies that included more than two manufacturers (N = 8, see above), only two studies had more than 42 subjects included, one of which is the one this power analysis is based on. The other, by Bruns et al., (2016) could have potentially performed a manufacturer comparison with enough statistical power.

**Figure 5.**
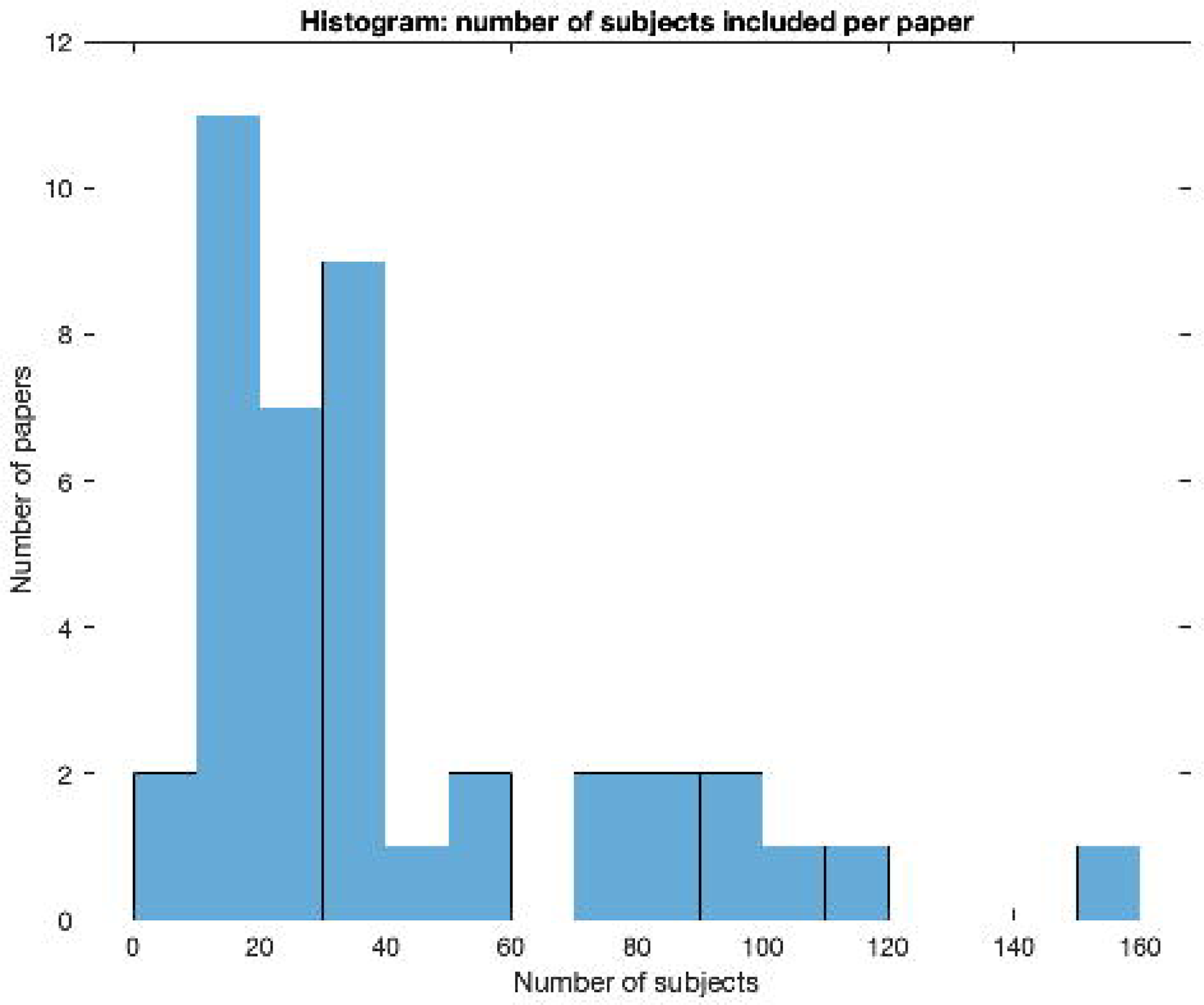
Distribution of the number of subjects with respect to the number of reviewed studies. Note that the outlier 2247 was removed for better visualization.

#### Pre-operative characteristics

##### Age

Mean or median age at implantation was reported in 83% of the studies (35 out of 42). Two studies reported only the age range. The mean age across articles was 56.8±9.9. The median was 59 (min: 25, max: 74). The distribution of mean/median age across studies is shown in Fig. 6.

**Figure 6.**
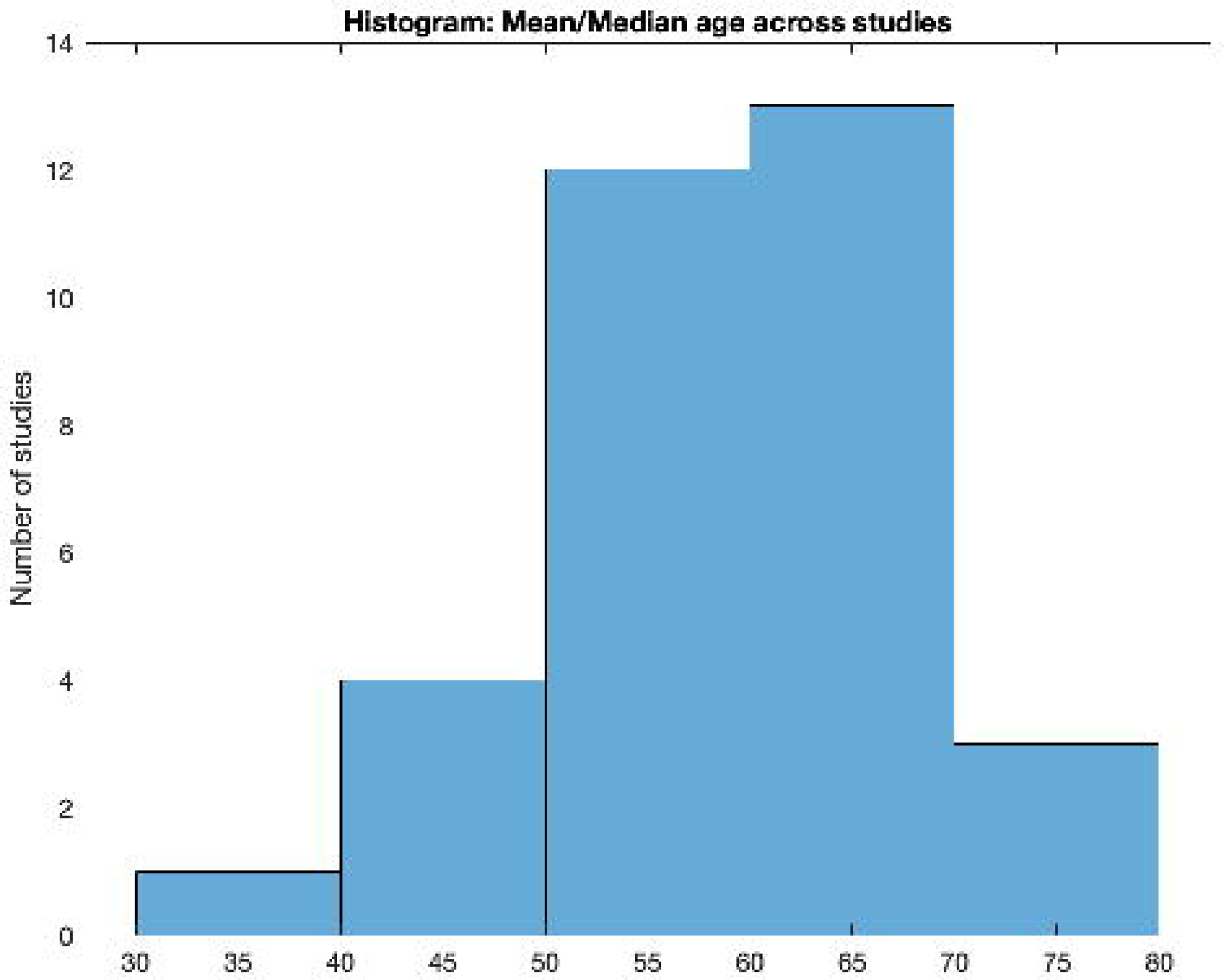
Mean/Median age distribution with respect to the number of reviewed studies

##### Etiology

The articles included in this review only reported on adults with postlingual hearing loss as per the exclusion criteria. Only 50% of the studies reported the etiology of hearing loss in the investigated cohort. Notably in all studies unknown etiology was also reported, these accounted in average for 46.9% of the patients. In Fig. 7, we show the number of studies per each etiology in the other 50%. Most studies mentioned a genetic/familial cause (N = 13). Next, Meningitis, Head trauma, Ototoxicity and Meniere’s disease were mentioned in 6-8 studies. Measles appeared in two studies only.

**Figure 7.**
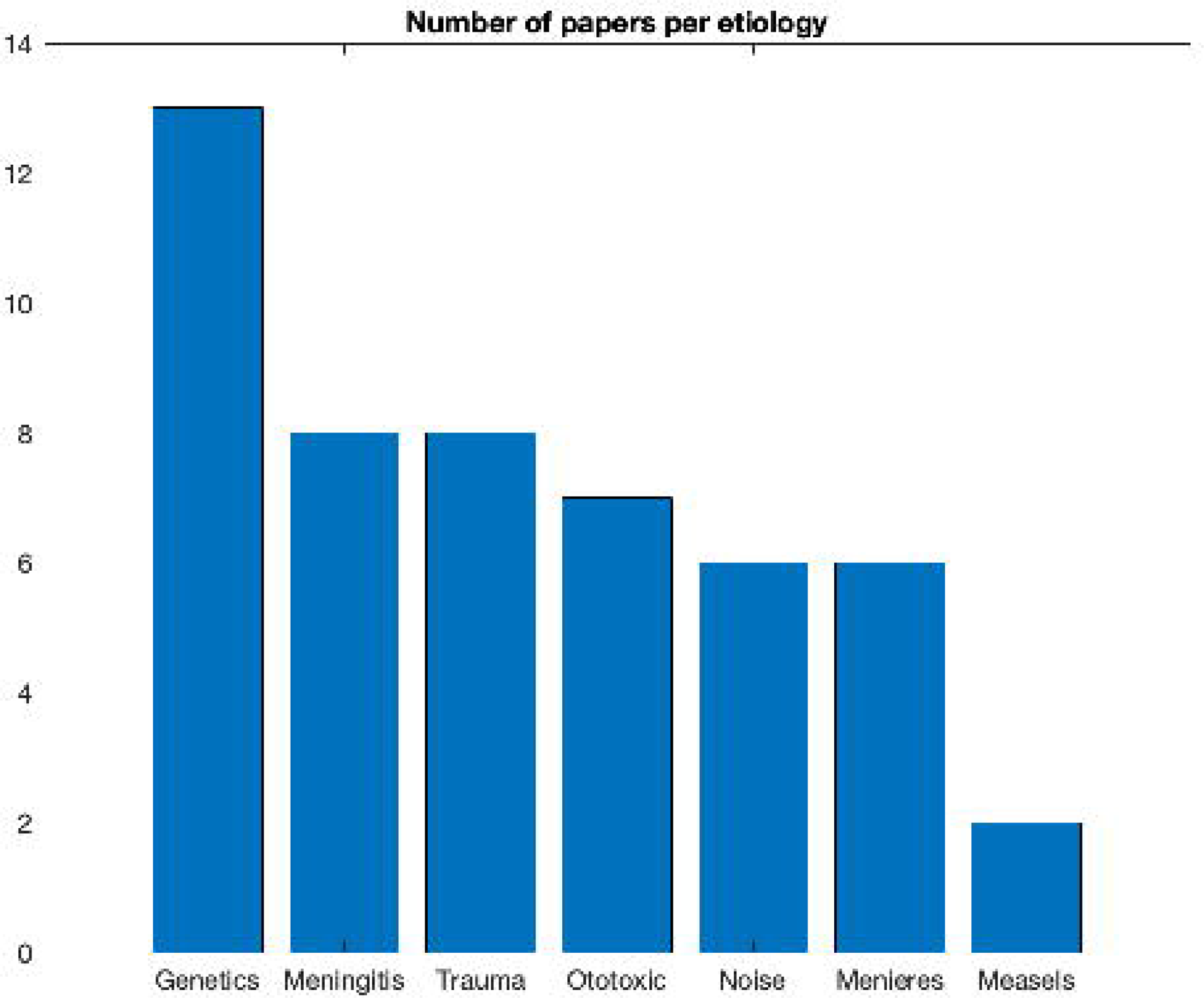
Distribution of etiologies with respect to the number of reviewed studies.

##### Duration of hearing loss

Only postlingual hearing loss was included in this scoping review. Results were obtained for each affected ear and not for a bilateral condition. Twenty out of 42 studies reported the duration of hearing loss or of deafness prior to implantation. Specifically, eight studies reported the duration of deafness, from which four reported the average (22.8, 3.2, 2.6, and 3.6) and three reported the range (0.3-41, 0.3-10, 1-6) of years with deafness prior to implantation across subjects. Twelve studies reported duration of hearing loss, from which, three studies reported the range in years of hearing loss across subjects (1-28, <10, <20) and nine studies reported the average in years of hearing loss across subjects (across studies: 22.2±8.7).

##### Hearing loss severity

Pre-operative average pure-tone detection was reported in 16 out of 42 studies in various forms, either directly stating in the text the average PTA or by supplying a pure-tone audiometric graphic from which we estimated the PTA at three-frequency (0.5, 1, 2 kHz) average. Note that depending on the graphic resolution, these values were not always accurate. Across these studies, we found that the average PTA was 89.5dB (STD 12.5).

#### Post-operative speech and music perception outcomes

Different types of speech and music performance tests were used to report post-operative outcomes. Thirty-three studies reported only speech outcomes, three studies performed both speech and music tests and nine studies reported only music tests. The assessment tool(s) used in publications pertaining only to music varied greatly. Six studies used surveys, questionnaires and Quality of Life measures to assess performance. Four studies used music samples, music bursts, instrument identification, timbre and intonation identification to assess outcomes. Two studies reported outcomes using validated music tests, specifically the Musical Sound Quality Impairments in Cochlear Implants (MUSHRA, Johns Hopkins University) and the Musical Sounds in Cochlear Implants perception test (MuSIC, Technical University Munich). Additional outcome assessment tools included objective measures and vocoded stimuli, i.e., synthesized signals that are thought to simulate, for a normal-hearing listener, the perception of speech as heard through a cochlear implant. In addition to a wide variety of outcome measures, we also found the aims of the studies were quite diverse, like the studies on speech perception outcomes, from assessing quality and enjoyment of sound to evaluating the impact of an online music training program. We found that most studies reported speech perception outcomes using the CNC word recognition test (N = 16), with AzBio, a sentence list test, mentioned in nine studies (see Fig. 8). In addition, 94% of all speech performance tests were conducted in noise conditions.

**Figure 8.**
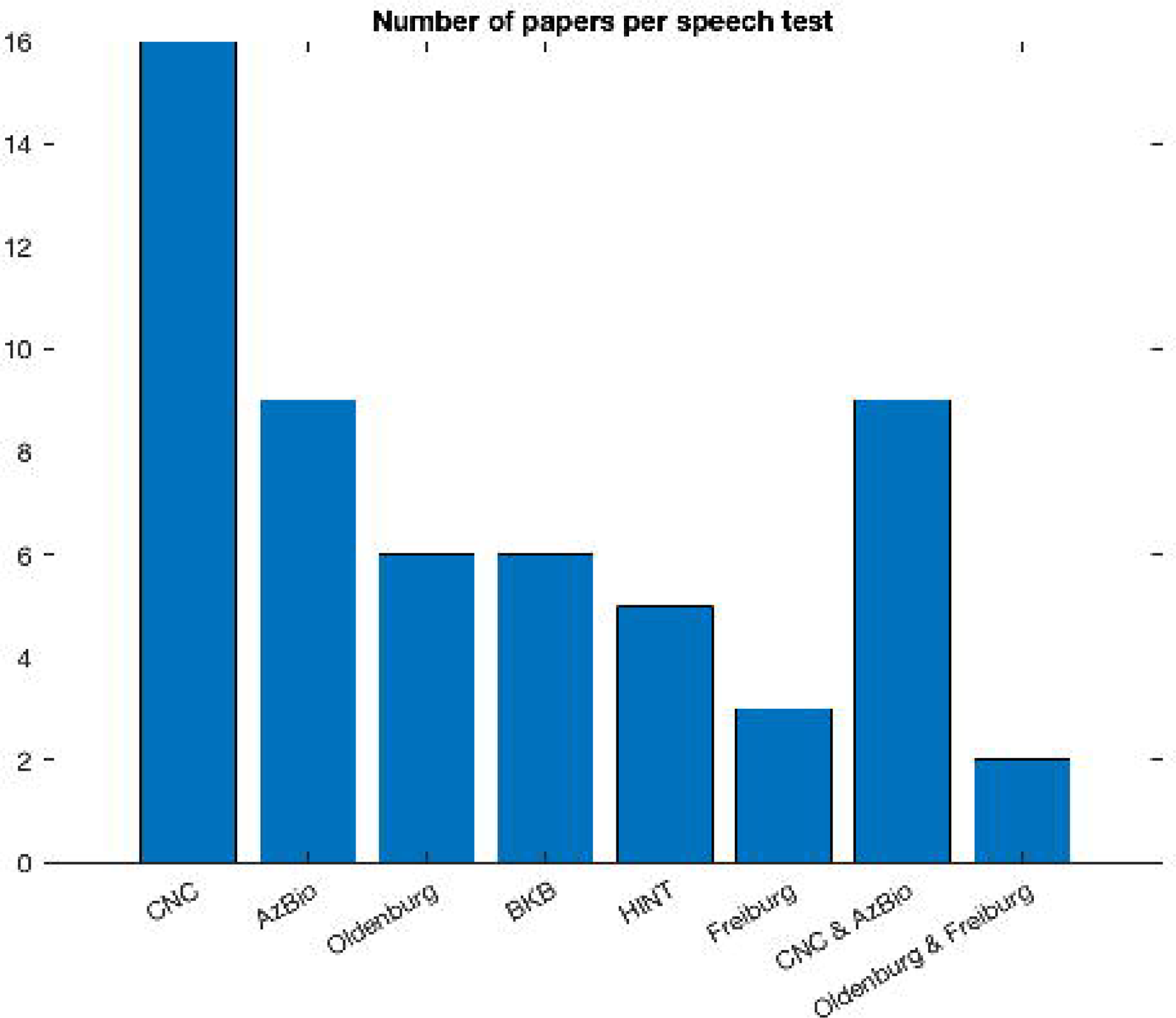
Distribution of speech tests with respect to the number of reviewed studies

Next, we specifically investigated the test conditions used within studies that reported on both CNC words and AzBio sentence tests. This was to assess whether results could be compared in a meta-analysis. We found that AzBio studies were performed adaptively and in each paper at a different presentation level (60 or 65 dB, with +5 or +10 dB SNR), which prohibited the possibility to summarize the scores.

On the other hand, studies reporting CNC test results had more commonalities. We identified six studies that reported the exact same conditions in which the test was conducted: non-adaptive 60dB SPL in quiet. From these studies, three studies were performed with CI systems by Cochlear and three studies with CI systems by MED-EL. In Table 1, we summarize the results reported for CNC in quiet in the respective studies. Note that some studies did not report the values directly in the text, and these needed to be extracted from the graphics, hence some small inaccuracies should be expected.

**Table 1:** Overview of publications that collected CNC word scores at a presentation level 60dB SPL in quiet.

| Paper | Manufacturer | Study location | N-subjects | 1-mo | 3-mo | 6-mo | 12-mo |
| --- | --- | --- | --- | --- | --- | --- | --- |
| Kelsall | Cochlear | Colorado, USA | 100 | - | 56% | 61% | 65% |
| Dillon (FSP) | MED-EL | N. Carolina, USA | 11 | 38% | 50% | 59% | - |
| Runge | Cochlear | Wisconsin, USA | 38 | - | 49% | 53% | 57% |
| Buchman | Cochlear | Missouri, USA | 96 | - | - | 61% | - |
| Buss | MED-EL | N. Carolina, USA | 20 | 39% | 43% | 59% | 57% |
| Canfarotta | MED-EL | N. Carolina, USA | 19 | 49% | 53% | 62% | 60% |

To account for the different number of participants in each study we performed a weighted averaging of the scores by the number of participants (see Table 2). We synthesized the results for different intervals including 1-month, 3-month, 6-month and 12-month following implant operation. Note that it was not possible to perform a meta-analysis of effect sizes in those studies, as some (Dillon, Buchmann, Buss, Canfarotta) did not report the necessary parameters (standard deviations – STD, standard errors of the mean - SEM). Notably, the results suggest that the differences between the CI-systems in terms of speech outcomes in quiet are negligible. Again, differences may exist when noisy or adaptive conditions are tested but data were not available to make such comparisons.

**Table 2:** Weighted averaging of speech scores across manufacturers

|  | 1-Mo | 3-Mo | 6-Mo | 12-Mo |
| --- | --- | --- | --- | --- |
| MED-EL | 42.6% | 48.3% | 60.1% | 58.5% |
| Cochlear | - | 54.1% | 59.7% | 62.8% |

## 4. Discussion

In this scoping review, we analyzed the literature for recent studies (2015 - 2022) reporting speech and music performance outcomes in adults implanted with a CI system. Our aim was: (1) to map the literature of speech and music performance outcomes and (2) to find whether studies perform outcome comparisons between devices of different manufacturers. Our findings show that very few publications directly compared patient performance outcomes between manufacturers of CI systems. Similar findings were recently reported by Stucke et al.^4^ for a limited selection of medical devices. CI device comparisons of different manufacturers are rare not only within an implant center but also across centers.

Possibly, the lack of performance-based manufacturer comparative publications is due in part to the fact that there is no consensus among large CI programs as to a systematic method of collecting outcome data. Differences in speech outcome measures (monosyllabic words, sentence in quiet, sentence in noise), differences in presentation levels (adaptive, fixed dB, various SNR’s) and inclusion of other measures such as quality of life (QoL) and music evaluation vary significantly between CI centers. Such differences make data collation a challenging task. For instance, Carlson et al. (2017) surveyed CI centers and found that 100% of responding clinics used AzBio sentences and 68% used speech in noise testing to determine candidacy. However, there was no consistency in the level of the noise used. Some centers report using a +10 SNR, others a +5 SNR and a majority used some combination of the two. As with pre-operative assessment for candidacy, a lack of consistency is seen when measuring post-operative outcomes.

In addition, performance-based manufacturer comparisons demand large numbers of implanted patients, to account for interindividual variability as well as other factors influencing performance outcomes (see above), which perhaps prohibit large CI centers from this task. Evidence suggests though, that large CI centers in Germany and USA perform speech perception tests as a routine procedure pre-implantation. For example, the FDA only approves clinical trials that use the CNC word recognition test as the primary measure for determining CI candidacy^5^. In addition, they recommend specific set-up for the tests that could be potentially adopted by large CI centers in the USA. In Germany the regulations are also specific. The Freiburger Test was recommended by the German society for otolaryngology in the publication „Weissbuch: Cochlea-Implantat (CI) - Versorgung in Deutschland“^6^. In accordance with this recommendation, a large CI center in Kiel, Germany, conducted retrospective analyses of speech performance outcomes using the Freiburger Test (Hey et al., 2016) in 626 persons implanted with a CI in the years 2010-2015. While most patients were implanted with a CI device by Cochlear (n = 165), some patients were also implanted by other manufacturers (MED-EL: n = 23, AB: n = 11). Despite the relatively low number of implants for comparisons between manufacturers, it is clear that data and methods are available for performing such meaningful comparisons, at least in Germany and the USA. Unfortunately, other countries such as France, Spain, Netherlands or the UK, have less-specific regulations regarding CI candidacy which subsequently leads to a strong variety in speech and music performance tests used^7^.

Another example that data and methods are available to perform comparisons is a study conducted by Kurz et al., (2019) in Wurzburg, Germany. In a retrospective data analysis of 55 subjects, the Freiburger Numbers, Freiburger Monosyllable and HSM sentences were examined at one, three, six and 12 months, as well as at yearly follow-up appointments. Similar to the Kiel study mentioned above, the number of implants was too low to compare between manufacturers, as they had only six subjects with either a Cochlear or AB device. Nonetheless, the presence of such data demonstrates that comparisons between device manufacturers are possible.

In the 42 publications that survived our selection criteria, we found that the vast majority mentioned the specific CI system manufacturer and 88% of those studies also mention the specific device used. In addition, we found six publications with >70 subjects, and one study included a sample of 150 subjects. With such large cohorts, inter-individual variability as well as other factors influencing performance may play a less significant role when averaging speech performance outcomes. Notably, sample sizes strongly varied between studies. We provided a power analysis for the comparison between CI systems of different manufacturers that assessed the minimum number of subjects required to perform meaningful manufacturer comparisons to be 42, when considering between-group comparisons.

In terms of outcome assessment, we found that most studies used speech performance tests to assess the beneficial effect of CI systems. Several studies have also used music performance tests to assess performance outcome following implantation. There was a strong variability in tests used to quantify speech and music performance post-operatively. Nonetheless, we identified 16 studies that used the CNC test under different conditions. From which, six studies that were conducted in the USA, which means that all subjects were English speakers, used similar CNC test conditions: non-adaptive 60dB SPL in quiet. This allowed us to perform a weighted average of CNC outcomes for different time intervals following implantation across the six studies. Note that summarizing the data across studies using effect-sizes was not possible since some of the studies did not report standard deviations or standard errors, which simply does not withstand good scientific practice. Notably, three studies featured MED-EL devices and three studies featured CI systems by Cochlear. The differences found were rather negligible and inconsistent, thus no statements could be made regarding superiority of one specific device. Although CNC in quiet is most commonly used in the studies reported here, sentence tests such as in AzBio are a better reflection of speech performance in real-life scenarios. However, as the words in the sentence are associated with each other, it is impossible to detach the effect of context on hearing ability (Gertjan Dingemanse & Goedegebure, 2019). In addition, the quiet condition is not valuable, since a far more important outcome is comprehension of speech in noise conditions. Unfortunately, studies implementing sentence tests (examples) in noise, varied significantly in terms of the specific test conditions, thus not allowing cross-studies comparisons. Optimally, post-operative comparisons between devices should be performed within one CI center. As stated above, there is evidence that CI centers in Germany routinely perform the Freiburger test post-operatively, at different time intervals. These data could be leveraged to fill the knowledge gap in terms of reliable manufacturer comparisons. Note however that we found only three studies that used the Freiburg test, probably as most reports on the Freiburg test are from German CI centers and in German and we excluded non-English journals.

Notably, in terms of music performance, we found extreme inconsistencies between studies: of the nine studies identified for in depth review, no two studies used the same test to assess outcomes. Some of the parameters tested were often similar but not identical to those in standardized tests such as MuSIC (Drennan et al., 2015; Drennan & Rubinstein, 2008; Gfeller et al., 2006). As for speech perception tests, music perception tests vary in various features. This prohibited us from performing any kind of summary analysis on these outcomes. Notably, music tests to assess outcomes of CI implantation are relatively new and not anchored in any reimbursement regulation and therefore are less reported and less consistent. As music performance is a significant measure that strongly affects patients’ quality of life (Dritsakis et al., 2017; Lassaletta et al., 2007), we urge future studies to follow a validated music test in future investigations. It would also be important to investigate how cultural differences affect CI users’ ability to perceive music. Indeed, previous studies could link between cultural aspects and different music perception parameters such as pitch discrimination, melody and rhythm (Wong et al., 2012; Zhang et al., 2020). These findings could assist with future collation of data across different CI centers around the world as well as guide CI manufacturers in adaptation of CI technology to achieve optimal music performance.

We tried to map the reasons regarding feasibility of comparative studies between CI systems of different manufacturers. Firstly, it would be important to compare technologies of the same generations. Large implant centers have patients using multiple generation devices which could make comparisons difficult. Secondly and as stated above, many subjects would be required to better control for inter-individual variability. Small to medium size centers may not have enough subjects to draw a comparison. Thirdly, and specifically related to cross-center comparisons, the evaluation measures, presentation levels, signal to noise ratios, test intervals and test language need to be consistent to make a valid comparison. In a retrospective study design, it is understandable that finding such consistency across centers is challenging. A prospective study would be easier to design but would also have its own challenges. Centers would need to implant the same generation technologies, match the subjects for age, duration of deafness or hearing loss and duration of implant use. Data collection would likely take several years to draw conclusive findings. Additionally, some patient related parameters that would be important to consider when comparing outcomes were either not reported or varied between studies. Long durations of deafness and certain etiologies are such examples that are associated with poorer outcomes and therefore should either be part of the exclusion criteria or matched pairs should be considered as they do not reflect the average population.

Although direct comparisons between manufacturers are scarce, we did find publications reporting performance outcomes in large numbers of subjects implanted with the same device. In these studies, data collection was very consistent and systematic. Patients were tested at defined test intervals using specific evaluation measures at the same presentation level and SNR. Potentially, large CI centers implanting sufficient number of devices (see power analysis above) from different manufactures could conduct retrospective comparisons using available data. Alternatively, if Center A with a large cohort reported on outcomes with device X and Center B, with an equally large cohort, reported on outcomes with device Y, a comparison between device outcomes could be made. Probably the most valid comparison of speech and music performance outcome of different CI manufactures, is in patients implanted with two different brands of CI (Harris et al. 2011). As mentioned in the introduction, most CI users implanted with two brands of CI, have an apparent preference towards one device, which clearly shows that necessity of providing comparative outcome data to the great benefit of the CI community.

### Limitations of this scoping review

Some limitations of this scoping review need to be mentioned. First, we used specific search terms “Cochlear implant outcomes adults” and “Cochlear implant music adults” which may have impacted the extent of studies found. Note however that an initial search with different terms have yielded similar results. The exclusion criteria for both patient indications and study types as well as publication language (English) may have also limited the number of publications available for review. Lastly, we restricted our search to recent publications from 2015 to be able to compare results of users with current generation technology. Reviewing literature published prior to 2015 would have produced more outcomes measures for comparison but with outdated technology.

## Conclusions

We found very few publications that compared speech and music outcomes across manufacturers. Performance data of different studies cannot be compared between manufacturers for various reasons, most notably being the variability in assessment measures and test conditions, as well as reporting bias. We argue that it is possible, however, to perform such quantitative comparisons. Data to do so should already exist at least in large CI centers in Germany and the USA but analyzes are not published despite the strong need by CI recipients and medical professionals. We therefore urge the community to make these data available. Future efforts should focus on forming a consensus regarding a systematic method of collecting outcome data following CI implantation which will serve clinicians and prospective CI recipients with performance-based comparisons between different CI systems and lead to an informed decision-making. In addition, this information could drive innovation in device design as well as future developments of CI systems that are focused on the patient’s experience. Lastly, systematically collected data could provide predictive information for clinicians regarding performance outcomes.

## Data Availability

All data produced in the present study are available upon reasonable request to the authors

https://github.com/Syte-Institute/Publications/blob/6e8c92d0a70589e1ceefb34ba1134143d2cf1de9/230613_CI%20outcomes%20speech%20and%20music.xlsx

## Acknowledgments

This study did not receive any funding. The authors state no conflict of interest. Syte Institute received financial compensation for consultation to Cochlear and MED-EL in the past.

## Availability of Data

The Excel spreadsheet can be accessed online: https://github.com/Syte-Institute/Publications/blob/6e8c92d0a70589e1ceefb34ba1134143d2cf1de9/230613_CI%20outcomes%20speech%20and%20music.xlsx

## Footnotes

1 https://blog.medel.com/changing-to-a-med-el-implant-is-the-best-thing-i-ever-did-enids-story/

2 https://www.medrxiv.org/content/10.1101/2022.05.30.22275158v1

3 www.cochlearimplanthelp.com

4 https://www.medrxiv.org/content/10.1101/2022.05.30.22275158v1

5 https://nap.nationalacademies.org/catalog/26057/evaluating-hearing-loss-for-individuals-with-cochlear-implants

6 https://cdn.hno.org/media/2021/ci-weissbuch-20-inkl-anlagen-datenblocke-und-zeitpunkte-datenerhebung-mit-logo-05-05-21.pdf

7 https://adulthearing.com/wp-content/uploads/2021/04/CIICA-Consensus-Statement.pdf

## References

Adunka, O. F., Gantz, B. J., Dunn, C., Gurgel, R. K., & Buchman, C. A. (2018). Minimum Reporting Standards for Adult Cochlear Implantation. Otolaryngology - Head and Neck Surgery (United States). https://doi.org/10.1177/0194599818764329

Ahmed Elkayal, V., Mourad, M., Elbanna, M., & Mohamed Talaat, M. (2016). Evaluation of factors that influence cochlear implant performance. Advanced Arab Academy of Audio-Vestibulogy Journal. https://doi.org/10.4103/2314-8667.191235

Assouly, K., Smit, A. L., Stegeman, I., Rhebergen, K. S., van Dijk, B., & Stokroos, R. (2021). Cochlear implantation for tinnitus in adults with bilateral hearing loss: protocol of a randomised controlled trial. BMJ Open, 11(5), e043288. https://doi.org/10.1136/BMJOPEN-2020-043288

Boisvert, I., Reis, M., Au, A., Cowan, R., & Dowell, R. C. (2020). Cochlear implantation outcomes in adults: A scoping review. In PLoS ONE. https://doi.org/10.1371/journal.pone.0232421

Bruns, E. A., el Haddad, I., Slowik, J. G., Kilic, D., Klein, F., Baltensperger, U., & Prévôt, A. S. H. (2016). Identification of significant precursor gases of secondary organic aerosols from residential wood combustion. Scientific Reports. https://doi.org/10.1038/srep27881

Carlson, M. L., Patel, N. S., Tombers, N. M., Dejong, M. D., Breneman, A. I., Neff, B. A., & Driscoll, C. L. W. (2017). Hearing Preservation in Pediatric Cochlear Implantation. Otology and Neurotology. https://doi.org/10.1097/MAO.0000000000001444

Carlson, M. L., Sladen, D. P., Gurgel, R. K., Tombers, N. M., Lohse, C. M., & Driscoll, C. L. (2018). Survey of the American Neurotology Society on Cochlear Implantation: Part 1, Candidacy Assessment and Expanding Indications. Otology and Neurotology, 39(1), e12– e19. https://doi.org/10.1097/MAO.0000000000001632

Chang, C. B. (2021). Phonetics and Phonology of Heritage Languages. In The Cambridge Handbook of Heritage Languages and Linguistics. https://doi.org/10.1017/9781108766340.027

Deshpande, P. R., Rajan, S., Sudeepthi, B. L., & Nazir, C. P. A. (2011). Patient-reported outcomes: A new era in clinical research. Perspectives in Clinical Research, 2(4), 137. https://doi.org/10.4103/2229-3485.86879

Dorman, M. F., & Gifford, R. H. (2017). Speech understanding in complex listening environments by listeners fit with cochlear implants. In Journal of Speech, Language, and Hearing Research. https://doi.org/10.1044/2017_JSLHR-H-17-0035

Drennan, W. R., Oleson, J. J., Gfeller, K., Crosson, J., Driscoll, V. D., Won, J. H., Anderson, E. S., & Rubinstein, J. T. (2015). Clinical evaluation of music perception, appraisal and experience in cochlear implant users. Http://Dx.Doi.Org/10.3109/14992027.2014.948219, 54(2), 114–123. https://doi.org/10.3109/14992027.2014.948219

Drennan, W. R., & Rubinstein, J. T. (2008). Music perception in cochlear implant users and its relationship with psychophysical capabilities. Journal of Rehabilitation Research and Development, 45(5), 779–790. https://doi.org/10.1682/JRRD.2007.08.0118

Dritsakis, G., van Besouw, R. M., & O’ Meara, A. (2017). Impact of music on the quality of life of cochlear implant users: a focus group study. Cochlear Implants International. https://doi.org/10.1080/14670100.2017.1303892

Dunn, C., Miller, S. E., Schafer, E. C., Silva, C., Gifford, R. H., & Grisel, J. J. (2020). Benefits of a hearing registry: Cochlear implant candidacy in quiet versus noise in 1,611 patients. American Journal of Audiology. https://doi.org/10.1044/2020_AJA-20-00055

Faul, F., Erdfelder, E., Lang, A. G., & Buchner, A. (2007). G*Power 3: A flexible statistical power analysis program for the social, behavioral, and biomedical sciences. Behavior Research Methods. https://doi.org/10.3758/BF03193146

Gertjan Dingemanse, J., & Goedegebure, A. (2019). The Important Role of Contextual Information in Speech Perception in Cochlear Implant Users and Its Consequences in Speech Tests. Trends in Hearing. https://doi.org/10.1177/2331216519838672

Gfeller, K. E., Olszewski, C., Turner, C., Gantz, B., & Oleson, J. (2006). Music Perception with Cochlear Implants and Residual Hearing. Audiology and Neurotology, 11(Suppl. 1), 12–15. https://doi.org/10.1159/000095608

Hey, M., Brademann, G., & Ambrosch, P. (2016). [The Freiburg monosyllable word test in postoperative cochlear implant diagnostics]. HNO.

Holden, L. K., Finley, C. C., Firszt, J. B., Holden, T. A., Brenner, C., Potts, L. G., Gotter, B. D., Vanderhoof, S. S., Mispagel, K., Heydebrand, G., & Skinner, M. W. (2013). Factors affecting open-set word recognition in adults with cochlear implants. Ear and Hearing. https://doi.org/10.1097/AUD.0b013e3182741aa7

Killan, C., Scally, A., Killan, E., Totten, C., & Raine, C. (2019). Factors affecting sound-source localization in children with simultaneous or sequential bilateral cochlear implants. Ear and Hearing. https://doi.org/10.1097/AUD.0000000000000666

Krueger, B., Joseph, G., Rost, U., Strau-Schier, A., Lenarz, T., & Buechner, A. (2008). Performance Groups in Adult Cochlear Implant Users. Otology & Neurotology. https://doi.org/10.1097/mao.0b013e318171972f

Kurz, A., Grubenbecher, M., Rak, K., Hagen, R., & Kühn, H. (2019). The impact of etiology and duration of deafness on speech perception outcomes in SSD patients. European Archives of Oto-Rhino-Laryngology. https://doi.org/10.1007/s00405-019-05644-w

Lassaletta, L., Castro, A., Bastarrica, M., Pérez-Mora, R., Madero, R., de Sarriá, J., & Gavilán, J. (2007). Does music perception have an impact on quality of life following cochlear implantation? Acta Oto-Laryngologica. https://doi.org/10.1080/00016480601002112

Lenarz, T., Zwartenkot, J. W., Stieger, C., Schwab, B., Mylanus, E. A. M., Caversaccio, M., Kompis, M., Snik, A. F. M., D’Hondt, C., & Mojallal, H. (2013). Multicenter study with a direct acoustic cochlear implant. Otology and Neurotology. https://doi.org/10.1097/MAO.0b013e318298aa76

Medicine, I. of M. (US) R. on E.-B., Yong, P. L., Saunders, R. S., & Olsen, L. (2010). Transparency of Cost and Performance. https://www.ncbi.nlm.nih.gov/books/NBK53921/

Munn, Z., Peters, M. D. J., Stern, C., Tufanaru, C., McArthur, A., & Aromataris, E. (2018). Systematic review or scoping review? Guidance for authors when choosing between a systematic or scoping review approach. BMC Medical Research Methodology, 18(1), 1–7. https://doi.org/10.1186/S12874-018-0611-X/TABLES/1

Neben, N., Buechner, A., Schuessler, M., & Lenarz, T. (2018). Outcome evaluation on cochlear implant users with residual hearing. Cochlear Implants International. https://doi.org/10.1080/14670100.2017.1390852

Shader, M. J., Nguyen, N., Cleary, M., Hertzano, R., Eisenman, D. J., Anderson, S., Gordon-Salant, S., & Goupell, M. J. (2020). Effect of Stimulation Rate on Speech Understanding in Older Cochlear-Implant Users. Ear and Hearing. https://doi.org/10.1097/AUD.0000000000000793

Sivonen, V., Willberg, T., Aarnisalo, A. A., & Dietz, A. (2020). The efficacy of microphone directionality in improving speech recognition in noise for three commercial cochlear-implant systems. Cochlear Implants International. https://doi.org/10.1080/14670100.2019.1701236

Spahr, A. J., Dorman, M. F., & Loiselle, L. H. (2007). Performance of patients using different cochlear implant systems: Effects of input dynamic range. Ear and Hearing. https://doi.org/10.1097/AUD.0b013e3180312607

Sturm, J. J., Patel, V., Dibelius, G., Kuhlmey, M., & Kim, A. H. (2021). Comparative Performance of Lateral Wall and Perimodiolar Cochlear Implant Arrays. Otology & Neurotology_J: Official Publication of the American Otological Society, American Neurotology Society [and] European Academy of Otology and Neurotology. https://doi.org/10.1097/MAO.0000000000002997

Varadarajan, V. v., Sydlowski, S. A., Li, M. M., Anne, S., & Adunka, O. F. (2020). Evolving Criteria for Adult and Pediatric Cochlear Implantation. Https://Doi.Org/10.1177/0145561320947258, 100(1), 31–37. https://doi.org/10.1177/0145561320947258

Varadarajan, V. v., Sydlowski, S. A., Li, M. M., Anne, S., & Adunka, O. F. (2021). Evolving Criteria for Adult and Pediatric Cochlear Implantation. In Ear, Nose and Throat Journal. https://doi.org/10.1177/0145561320947258

Williges, B., Wesarg, T., Jung, L., Geven, L. I., Radeloff, A., & Jürgens, T. (2019). Spatial Speech-in-Noise Performance in Bimodal and Single-Sided Deaf Cochlear Implant Users. Trends in Hearing. https://doi.org/10.1177/2331216519858311

Wong, P. C. M., Ciocca, V., Chan, A. H. D., Ha, L. Y. Y., Tan, L. H., & Peretz, I. (2012). Effects of culture on musical pitch perception. PLoS ONE. https://doi.org/10.1371/journal.pone.0033424

Zeng, F.-G. (2022). Celebrating the one millionth cochlear implanta). JASA Express Letters, 2(7), 077201. https://doi.org/10.1121/10.0012825

Zhang, L., Xie, S., Li, Y., Shu, H., & Zhang, Y. (2020). Perception of musical melody and rhythm as influenced by native language experience. The Journal of the Acoustical Society of America. https://doi.org/10.1121/10.0001179

